# Effectiveness, facilitators and barriers of digital mental health services for First Nations Peoples in Australia: A systematic review

**DOI:** 10.1101/2025.04.27.25326251

**Authors:** Siyu Zhai, Andrew Goodman, Anthony C Smith, Sandra Diminic, Xiaoyun Zhou

## Abstract

**Background:** First Nations Peoples in Australia have unique health views. However, due to colonisation and intergenerational trauma, despite the strengths they have, the health inequity in mental health that First Nations Peoples are experiencing is still significant. These historical factors, combined with geographical remoteness and limited access to culturally safe services, resulted in mental health service gaps. Digital mental health (DMH) services, which offer interventions through digital platforms, are considered potential solutions to address this gap. However, there is limited evidence on the effectiveness of DMH for First Nations Peoples in Australia.

**Aim and Objectives:** This systematic review aimed to assess the effectiveness of DMH services in improving mental health outcomes for First Nations Peoples in Australia and to identify the facilitators and barriers that influence the implementation of DMH services in this context.

**Methods:** A systematic search was conducted across six academic databases to search for studies related to DMH services for First Nations Peoples in Australia. Search terms relating to First Nations Peoples, geographic terminologies of Australia, mental health, specific mental health conditions and digital mental health services were used. Studies were included if they assessed the effectiveness of digital mental health interventions among First Nations people in Australia. Effectiveness was defined as the ability of the DMH service to achieve its major targeted mental health outcome(s) in the current study. The data were extracted based on its study design, targeted services, and research findings, then synthesised using a thematic analysis framework.

**Results:** In total, 22 studies met the inclusion criteria. The included studies used a variety of study designs and researched multiple DMH services designed to provide support, treatment, and psychological assessments. A general effectiveness for non-severe mental health conditions was observed in the included studies. Several determinants of facilitators and barriers of the implementation of DMH services were identified, including: 1. Organisational and administrative factors; 2. Cultural appropriateness; 3. Accessibility; 4. Integration of DMH services to the existing situation; 5. Engagement between clients and service providers; 6. Coverage of different conditions and clients; 7. Acceptability to DMH services; 8. Digital literacy, and 9. Efficiency.

**Conclusion:** Given the effectiveness in providing services to most mental health conditions, DMH services have the potential to address the mental health needs of First Nations Peoples in Australia. However, the decision-making at multiple layers, as well as the design and implementation of DMH, should consider the determinants identified by this review.

## Introduction

Mental health is a state that helps people maintain mental well-being, realise their abilities, cope with life stresses and be free from clinically significant mental disturbances (1). Aboriginal and Torres Islander peoples in Australia (In this paper, the term ‘First Nations Peoples’ is respectfully used) have a collective culture and view mental health as a part of ‘Social and Emotional Well-Being’ that represents a more holistic view of health across all life stages (2, 3). Moreover, maintaining a personal connection to traditional culture, community, and lands serves as an important protective factor for First Nations Peoples’ mental health (2, 4–7) by reinforcing cultural strength and resilience, which would, in turn, foster personal resilience and self-esteem, thereby protecting their individual mental health (8) and reducing the likelihood of developing mental illness (6, 9, 10). When treating existing mental illness, the mental health services that acknowledged the right of self-determination and the need for cultural understanding of First Nations Peoples (11), provided culturally appropriate care (12) or employed First Nations,· staff and health workers (13) would also reinforce protective factors and achieve better treatment outcomes and patient satisfaction.

However, First Nations Peoples are facing a more complex mental health challenges than the general population in Australia due to external historical and social determinants (14). Historically, colonisation led to the loss of sovereignty, lands and customary law, which resulted in intergenerational trauma that worsened their overall social and emotional well-being and mental health (15). Moreover, traumatic child welfare policies, including “the stolen generation”, which forcibly separated First Nations Children from their families, further exacerbated trauma within the communities, then adversely affected the mental health of First Nations parents and children (10). Currently, First Nations Peoples continue to face daily discrimination, contributing to poorer mental health outcomes (16), along with inequity in education, employment, income, housing and other social disadvantages (10). These social determinants, coupled with structural inequities, increase the likelihood of mental health issues within these communities (17) and create mental health outcomes gaps between First Nations people and the general population.

For instance, the national health survey in 2014-2015 shows the prevalence of any mental illness among First Nations Peoples was 29.3% (9). Although this prevalence was reduced to 24% in 2018-2019, it was still considered high (18). Additionally, according to data collected from regional samples but applied to structured clinical interviews in 2014-2016, the prevalence of any mental illness was 42.2%, which is 4.2-fold higher than the general population in Australia (6). Another study involving clinical diagnosis and regional samples shows the lifetime prevalence of any mental illness of First Nations Peoples is 18.3%, significantly higher than the 5.7% prevalence of non-First Nations Peoples (19). According to an estimation based on eight nationally representative population surveys in Australia, the prevalence of anxiety and mood disorders among First Nations Peoples was 1.6-3.3 times the national prevalence (20). Furthermore, Compared to 6% of deaths accounted to suicide for the general population, 16% of deaths of First Nations Peoples were accounted to suicide in 2001-2005 (5).

Despite the significant mental health needs and gaps, First Nations Peoples also experience barriers to accessing mental health services. Firstly, the stigma towards mental health conditions, low health literacy and less comfort in seeking help from health professionals prevent First Nations people from using more mental health services (12, 13, 21). Secondly, the structural racism and discrimination against First Nations Peoples led to low utilisation of mainstream mental health services (13). Thirdly, the lack of local services creates barriers to travel for First Nations Peoples (12), especially for areas with higher geographical remoteness that have a higher proportion of First Nations populations (22) but lower service access (23) due to the lack of services and service providers (24). Also, culturally safe services may be unavailable for some First Nations Peoples, such as a lack of community-controlled mental health services (25) or health workers from non-First Nations Backgrounds not receiving appropriate cultural safety training to provide services (24, 26–28). As a consequence, in 2018-2019, only 31% of First Nations Adults with high or very high levels of psychological distress received services from mental health professionals (29). Moreover, mental health and well-being were the areas that mostly reported having health service gaps by the First Nations primary health organisations (134 out of 198 organisations) (30), indicating significant health challenges. Moreover, this health challenge faced by First Nations Peoples was disproportional compared to non-First Nations population in Australia. For instance, Among suicide cases, the First Nations population were approximately two times less than the non-First Nation population to have received mental health support (23.8% vs 43.3%) (31).In the Northern Territory, the service utilisation of First Nations Youth (aged under 25 years old) is nearly half of that of the non-First Nations Youth (1.9% versus 4.1%) (32).

Digital mental health (DMH) services refer to providing mental health services through digital technologies (33–35). DMH services can be implemented for the prevention, screening, intervention, and rehabilitation of mental health conditions (36) through mental health professionals or guide clients to self-help (37). A wide range of mental health conditions could be covered by DMH services, including anxiety disorders (38), depressive disorders (39) and post-traumatic stress disorder (40) or providing alcohol and other drug treatment supports (41). Also, DMH services can be delivered to different populations, including children and young people (42), adults (43) or older populations (44). In conclusion, digital mental health services offer a flexible and scalable approach to delivering mental health care across diverse service types, conditions, and populations.

DMH services have been applied to First Nations people worldwide (45–48) and show the potential to address current health outcome gaps faced by First Nations people in Australia. Global evidence from Canada, New Zealand and the United States shows DMH services could provide improvements comparable to existing face-to-face services for First Nations Peoples in the psychological assessment area (49–52). After applying DMH services in treatment and embedding DMH into general mental health services, positive clinical outcomes (51, 53) and higher continuity in receiving treatments (54) were found.

International evidence suggests DMH services can reduce the mental health gaps by improving the accessibility and availability of mental health services faced by First Nations Peoples (23, 34–36). Although First Nations Peoples in Australia face digital gaps compared to the general population, which hinder the implementation of DMH services (55), necessary digital infrastructure was started to be provided by governmental funding for rural and remote First Nation Peoples communities in Australia (56), and made addressing the digital gap possible. Another service gap faced by First Nations Peoples is the need for culturally appropriate services (25). Evidence shows digital health services still face barriers in addressing this gap, either by failing to provide such services (57, 58) or due to First Nations Peoples’ preference for the familiarity and safety of face-to-face services (52). However, some First Nations peoples have reported that digital health can facilitate the provision of culturally appropriate healthcare by increasing the involvement of First Nations Health Workers (45, 59) and some patients feel satisfied by seeing familiar faces through video conferencing (56). Such familiarity can also be provided by telehealth by reducing the need for travel and then facilitating the community and family members to join in the group decision-making (56). Besides, a systematic review reported the design of selected DMH services for First Nations Peoples in Australia were found to involve the community-led design to ensure cultural safety (60). Therefore, despite the evidence being mixed and uncertain, DMH holds the potential to address the gap in culturally safe mental health services.

Currently, most systematic reviews focus on international evidence on applying DMH services to First Nations peoples (45–48), and existing Australian evidence is more likely to review the broad and general health conditions rather than focus on digital mental health (49, 56, 59, 61) or solely focus on the design and assessment of DMH services for First Nations Peoples, rather than the implementation (60). There is a paucity of evidence on the effectiveness of DMH for First Nations people in Australia. However, such studies are scarce. Our study aims to address this research gap by exploring the following research questions: (1) What is the effectiveness of DMH services applied for First Nations Peoples Australia? (2) What are the determinants (i.e., facilitators and barriers) that affect First Nations people’s use of DMH? Such evidence is critical for health providers, policymakers and First Nations users to make evidence-informed decisions while prescribing, implementing and choosing DMHs.

## Methods

### 1. Defining determinants

In this study, the determinants include the facilitators, barriers and effectiveness. The facilitators and barriers are defined as determinants that can promote and impede the implementation of DMH services for First Nations Peoples in Australia. The effectiveness of DMH is defined as the extent to which DMH services obtain their desired outcomes. For example, DMH services which are related to psychological diagnosis and assessment (51), the accuracy of diagnosis is considered as effectiveness. While treatments provided through DMH services (53, 54, 62), the effectiveness is defined as the degree to which a treatment service achieves its intended therapeutic outcomes.

### 2. Searching Strategy

A systematic search was conducted during September 2024 across six online databases: PsycINFO, PubMed, Medline, Embase, Web of Science, and Google Scholar, with the results from the latter limited to the first 30 pages.

Studies related to the determinants of DMH services for First Nations Peoples in Australia were included. (See Appendix A for detailed inclusion/exclusion criteria and PICO protocol.)

Boolean operators, truncations, and search terms were tailored to each database. The search terms were organised around five themes: First Nations Peoples, geographic terminologies of Australia, mental health (including specific mental health conditions) and digital mental health services. (See Supplementary File S1 for details of searching strategies.)

### 2. Study Selection

Selected studies were imported into EndNote 20 and Covidence, and duplicates (N = 536) were removed at this stage. Afterwards, the titles and abstracts were screened. Studies irrelevant to this study topic (N = 712) in the title and/or abstract were excluded. Studies that had insufficient information in the title and abstract were moved to the full-text screening. During the full-text screening, articles that matched the exclusion criteria (N = 107) were excluded (See Figure 1 for PRISMA flow chart).

One reviewer (SZ) conducted the title and abstract screening; however, uncertainty in this process was solved by the discussion with another reviewer (XZ), and any disagreements were resolved by involving the third reviewer (SD), while two reviewers finished the full-text screening where one reviewer (SZ) made initial decisions and the other reviewer (XZ) checked. Therefore, the selection process was not blinded. The disagreements were solved by discussion or involving the third reviewer (SD).

### 3. Data extraction

Extracted data were tabulated, including author, year of publication, methods applied, study type (qualitative, quantitative or mixed methods), study aim, study population, research setting, relevant type of digital mental health intervention, relevant outcome (facilitators, barriers or effectiveness), and key research findings. (See Supplementary File S2 for the details of extracted data.)

For missing data, if the author did not specify the reason for the data missing, the reviewers tried to contact the author to obtain relevant information. If such information were unavailable or no response was received, the missing data would be excluded from the analysis process.

### 4. The strategy of data synthesis

The synthesis of data followed the thematic analysis method introduced by Thomas and Harden (63). The selected articles were coded into descriptive themes; then, the reviewers generated new analytical themes based on descriptive themes to present the primary results.

### 5. The Quality assessment

This study applied the Aboriginal and Torres Strait Islander Quality Appraisal Tool (64) to assess the quality of health research from a cultural sensitivity perspective. Overall, the included studies presented an acceptable connection to First Nations values and principles, except for a study that researched DMH services designed for the general population (65) and studies applied quantitative methods that resulted in lower participation in First Nations communities (66–68). Also, the current review applied the Mixed Methods Appraisal Tool (69) for assessing the methodological quality of included studies. Notably, only the relevant part of the included study would be assessed. For instance, if the qualitative part of a mixed-methods study is the only part that is relevant to the current review, the study would be appraised as a qualitative study. See Supplementary File S3 and S4 for the quality assessment results.

## Results

### 1. General characteristics of the studies

A total of 22 articles were included in this systematic review (65–68, 70–87). (See supplementary file S2 for detailed characteristics). Except for one study that only reported barriers (65) and four studies only reported the effectiveness (66, 68, 76, 86), the rest of the studies reported more than one targeted determinant (i.e. facilitators, barriers and effectiveness) of the current study. Among the included studies, there were eight qualitative studies (70, 71, 73, 77, 78, 81, 82, 87), eight mixed-method studies (72, 74, 75, 79, 80, 83–85), and six quantitative studies (65–68, 76, 86).

All studies involved First Nations Peoples as the clients of DMH services. There were five studies conducted on healthcare providers who provided DMH services to the First Nations Peoples (70, 73, 81–83), two studies involved both clients and service providers (71, 77), twelve studies recruited adult clients or clients whose ages were unspecified (65–68, 72, 74–76, 78, 84–86), three studies recruited youth or student clients (79, 80, 87).

#### 1.1. Targeted mental health conditions

Five studies that did not specify the mental health conditions targeted by DMH services (70, 71, 74, 80–83). Three studies focused on psychological assessment and diagnosis (66–68), while five studies addressed the management of distress, depression and anxiety disorders (67, 78, 85, 86, 88). Additionally, five studies examined the management of substance use (75, 76, 78, 79, 87), and another five were related to the management of suicide or self-harm behaviours (65, 71, 78, 85, 86).See Table 1 for detailed information about the setting of included studies.

**Table 1.** Settings of included studies.

| Author, Year | Study Aim | Participants | Research Setting | Digital Mental Health Service Type | Targeted mental health issue |
| --- | --- | --- | --- | --- | --- |
| Bennett-Levy et al. 2017 | Report the barriers and enablers of e-MH uptake in health providers who provide services to First Nations Peoples. | 50 providers of different mental health services for First Nations Peoples. | Two locations (Lismore and Tweed Heads) in northern New South Wales | General e-Mental Health (eMH) services (did not specify service type) | Unspecified |
| Brown et al. 2020 | Understand how mobile apps support suicide prevention gatekeepers in First Nations communities. | 12 participants (Indigenous health workers or community members) | A regional city in South West Queensland called Toowoomba | Suicide prevention efforts supported by mobile apps | Suicide and self-harm behaviours |
| Dingwall et al. 2023 | Assess the feasibility, acceptability, and use of the AIMhi-Y app and determine the feasibility of study procedures for future assessments. | 30 First Nations young people aged 12-25 years. | Darwin, Northern Territory (NT) | Aboriginal and Islander Mental Health Initiative for Youth (AIMhi-Y) app | Unspecified |
| Dingwall et al. 2015 | To develop and determine the acceptability, feasibility, and appropriateness of the Australian Integrated Mental Health Initiative [AIMhi] Stay Strong App for service providers working with First Nations Peoples. | 15 service providers, including health professionals, managers, programme coordinators, and an Aboriginal elder | Rural and remote primary health care service settings in the Northern Territory | The Australian Integrated Mental Health Initiative (AIMhi) Stay Strong App | General mental health area (unspecified) |
| Fletcher et al. 2017 | 1. To test the acceptability and feasibility of developing the "Stayin' on Track" website, which offers tailored support and information to young Aboriginal fathers.<br>2. To adapt and test the mobile phone-based text-messaging and mood-tracker program (SMS4dads program) that provide mental health supports to young Aboriginal fathers. | 20 Young Aboriginal fathers in the test of the "SMS4dads" smartphone program, 20 Young Aboriginal fathers and an uncertain number of community members in the yarn-up session on the "Stayin' on Track" website. | One regional city and two rural towns in New South Wales | "SMS4dads" smartphone program<br>"Stayin' on Track" website | General mental health area (unspecified) |
| Kennedy et al. 2021 | Describe the development (Stages 1 and 2) and pretest (Stage 3) of a prototype multi-behavioural change app MAMA-EMPOWER. | Stage 1: 8 First Nations Women<br>Stage 2: 6 First Nations Women<br>Stage 3: 16 First Nations women | Newcastle and Tamworth, New South Wales | Multibehavioural change app "MAMA-EMPOWER" | Substance use and psychological distress |
| Lee et al. 2019 | Validation of a survey app (Grog Survey App) that explores the alcohol consumption status among First Nations Peoples | 20 non-drinkers, 40 non-dependent drinkers and 40 dependent drinkers who are self-identified as First Nations Peoples and aged 16+ years old. | An Indigenous primary health care service and surrounding community in urban Queensland, and a regional ACCHS and a remote Aboriginal community-controlled drug and alcohol day centre (a drop-in service) in South Australia. | Grog Survey App | Substance use (Alcohol consumption) |
| Lee et al. 2019 | Validation of a survey app (Grog Survey App) that explores the alcohol consumption status among First Nations Peoples | 20 non-drinkers, 40 non-dependent drinkers and 40 dependent drinkers who are self-identified as First Nations Peoples and aged 16+ years old. | An Indigenous primary health care service and surrounding community in urban Queensland, and a regional ACCHS and a remote Aboriginal community-controlled drug and alcohol day centre (a drop-in service) in South Australia. | Grog Survey App | Substance use (Alcohol consumption) |
| Ma et al. 2022 | To understand the expectations of different communities to telehealth crisis support services in Australia. | 1300 Adults living in Australia, Including 2.4% First Nations participants | National setting in Australia | Lifeline Australia | Suicide and self-harm behaviours |
| Perdacher et al. 2022 | To determine the feasibility of the Stay Strong app as a digital well-being and mental health tool for use by First Nations Peoples in prison. | 27 clients and 10 health practitioners | 3 high-security prisons in Queensland, Australia | Stay Strong App (custodial version) | General mental health area (unspecified) |
| Povey et al. 2016 | To explore First Nations community members' experiences of using two e-mental health apps and identify factors that influence the acceptability. | 9 First Nations adults | Darwin, Northern Territory (NT) | AIMhi Stay Strong iPad app and the ibobbly suicide prevention app | AIMhi Stay Strong app: General mental health issues and substance use<br>ibobbly app: Psychological distress, depressive symptoms, suicide prevention |
| Povey et al. 2022 | To present an in-depth account of the second phase of participatory design in the development of the Aboriginal and Islander Mental Health Initiative for Youth (AIMhi-Y) app. | 75 First Nations youth | Across Australia | AIMhi-Y app (under design) | General mental health area (unspecified) |
| Povey et al. 2020 | Reports the result of the formative stage of the AIMhi-Y App development process which engaged First Nations youth in the co-design of the new culturally informed AIMhi-Y App. | 45 First Nations youth | Northern Territory (NT), Australia | General e-Mental Health (eMH) services (did not specify service type) | General mental health area (unspecified) |
| Puszka et al. 2016 | to understand stakeholder perspectives on the requirements for implementing DMH services in regional and remote health services for First Nations Peoples. | 32 stakeholders who provide mental health services for First Nations Peoples | Northern Territory of Australia | General e-Mental Health (eMH) services (did not specify service type) | general mental health area (unspecified) |
| Raphiphatthana et al. 2020a | To evaluate the process and the effectiveness of the e-mental health program. | 66 participants from First Nations primary care organisations | Remote Northern Territory communities, Darwin, Alice Springs, and Adelaide. | Did not specified, but most participants used Stay Strong App | General mental health area (unspecified) |
| Raphiphatthana et al. 2020b | This study aimed to understand service providers' perspectives and experiences of electronic mental health adoption. | 57 service providers working with Aboriginal and Torres Strait Islander people who had undergone electronic mental health training workshops. | Darwin, Alice Springs, and remote Northern Territory communities | General DMH services (but many of the responses were related to the use of the Stay Strong app) | Unspecified |
| Routledge et al. 2022 | To assess the acceptability and feasibility of Strong & Deadly Futures, a culturally inclusive alcohol and drug prevention program for First Nations secondary students. | 19 First Nations students out of 281 students | Two Independent urban schools and two rural state schools | Strong & Deadly Futures, a six-lesson, web-based package combining online illustrated storylines with interactive classroom activities | Substance use (Both alcohol and tobacco) |
| Russell et al. 2021 | To examine the utility of using a culturally appropriate dementia screening tool (KICA-screen) in a telehealth setting. | 33 medically stable First Nations inpatients/outpatients | Two local rural healthcare settings | Kimberley Indigenous Cognitive Assessment (KICA-screen) delivered via telehealth | Cognitive assessment |
| Snijder et al., 2021 | To describe the development of Strong & Deadly Futures, a web-based substance use education class. | 41 First Nations students and 36 non-First Nations students | Four schools in New South Wales | Strong & Deadly Futures, a six-lesson, web-based package combining online illustrated storylines with interactive classroom activities | Substance use (Both alcohol and tobacco) |
| Tighe et al. 2017 | To evaluate the effectiveness of a self-help mobile app (ibobbly) targeting suicidal ideation, depression, psychological distress and impulsivity among Indigenous youth in remote Australia. | 61 First Nations Australians aged 18-35 years | Remote and very remote communities in the Kimberley region of North Western Australia. | ibobbly programme | Suicidal ideation, depression, psychological distress and impulsivity |
| Tighe et al. 2020 | To discover pilot usage and acceptability data from the iBobbly suicide prevention app, an app distributed through a randomised controlled trial. | 13 First Nations Australians aged 18-35 years | Remote and very remote communities in the Kimberley region of North Western Australia. | ibobbly programme | Suicidal ideation, depression, psychological distress and impulsivity |
| Titov et al. 2019 | To report on Aboriginal and Torres Strait Islander (Indigenous) users of MindSpot, a national service for the remote assessment and treatment of anxiety and depression. | 780 First Nations participants | National setting across Australia | MindSpot service, including diagnosis and treatment courses | General mental health area, but this study focused on psychological distress, depression and anxiety. |
| Veinovic et al. 2023 | To evaluate mental state examinations that were delivered face-to-face and their telehealth version (MMSE and KICA-screen, TMMSE and TKICA-screen) among older First Nations Peoples. | 20 First Nations Peoples aged 55-69 years | Urban and regional New South Wales, Australia. | Telehealth version of Mini Mental State Examination (TMMSE) and Kimberly Indigenous Cognitive Examination short form (TKICA-screen) | Cognitive assessment |

### 2. Effectiveness

Twelve studies (66–68, 72, 75–78, 83–86) reported effectiveness of DMH services in managing mental health conditions among First Nations People in Australia (Table 2).

**Table 2.**
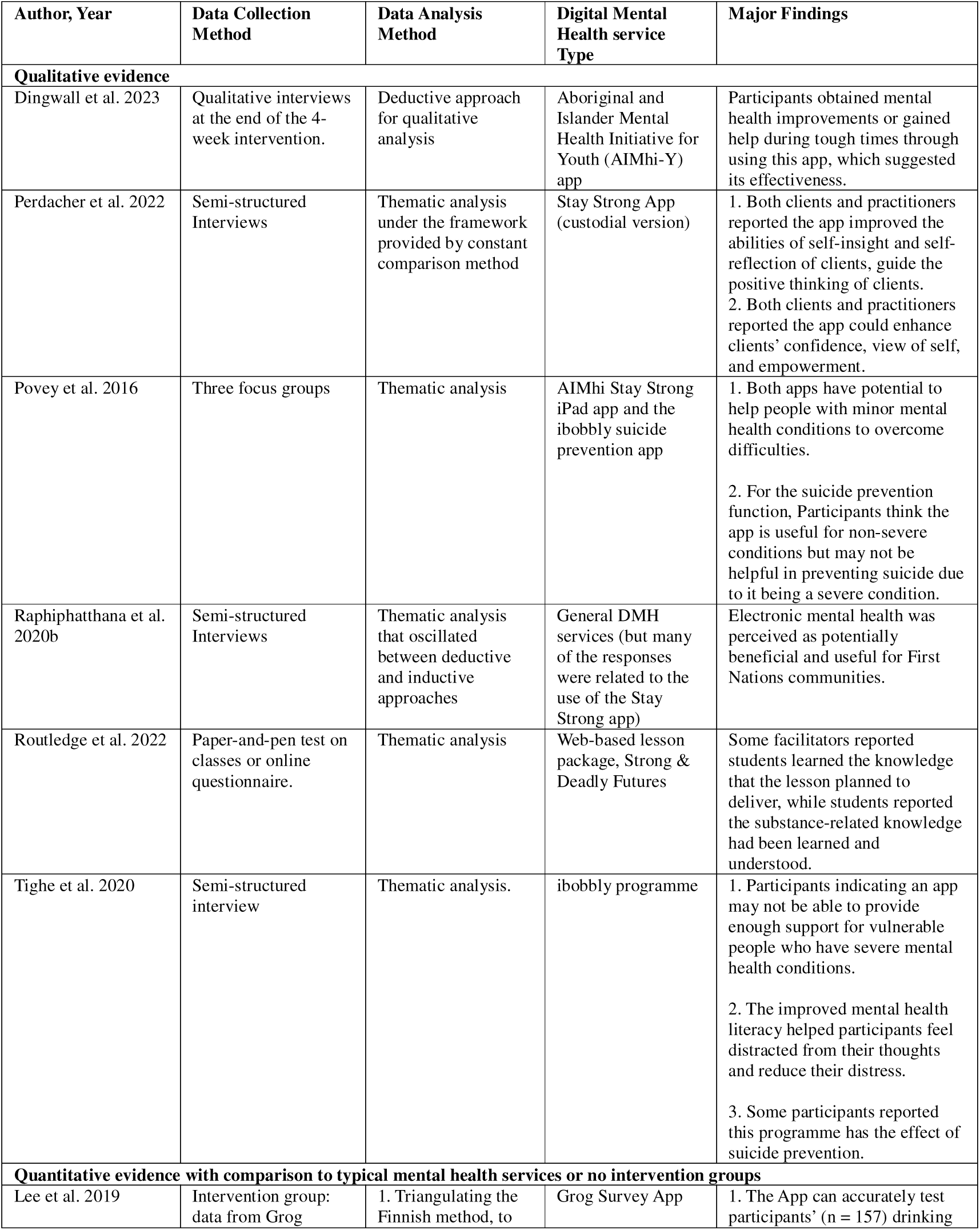
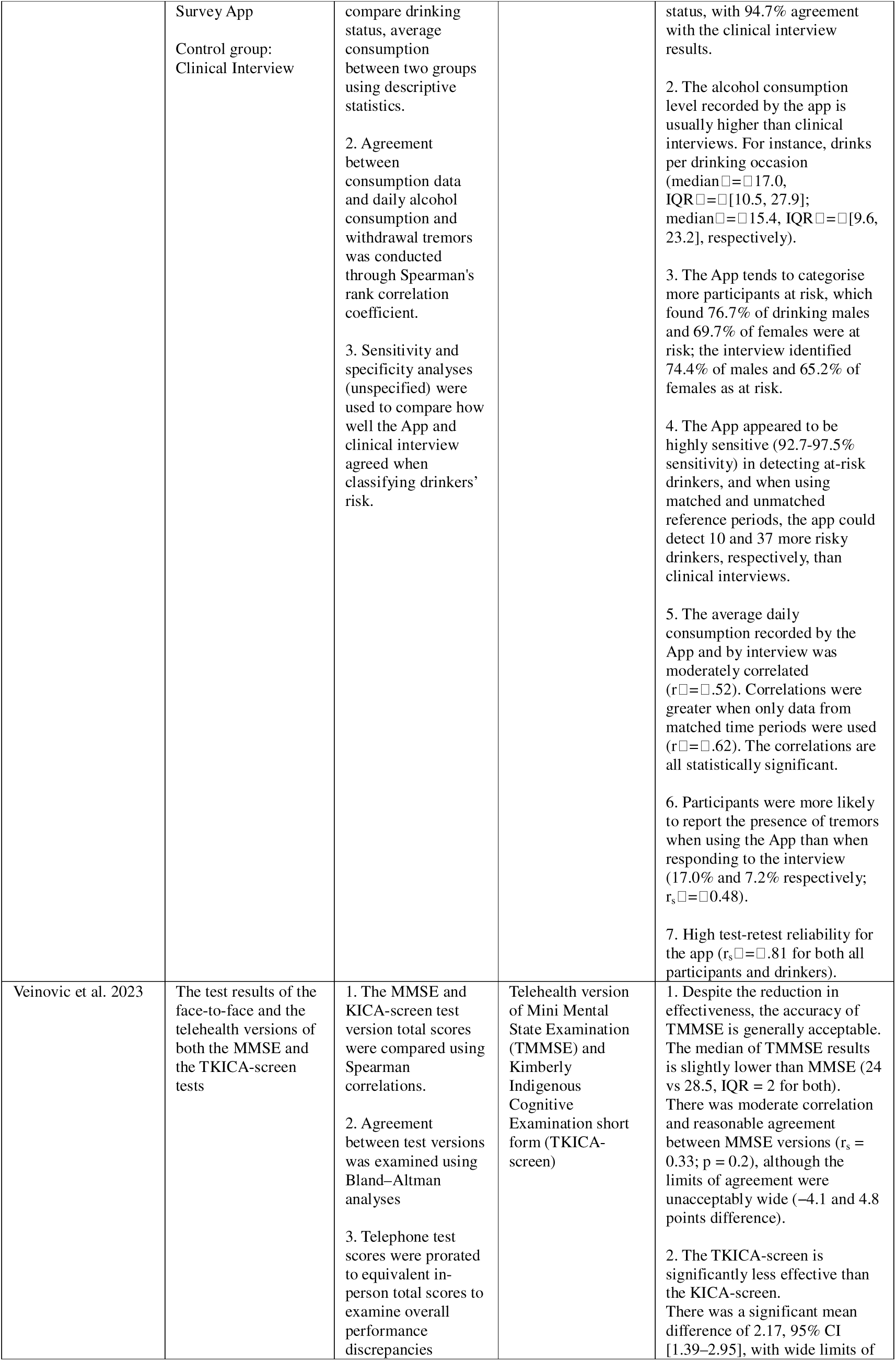
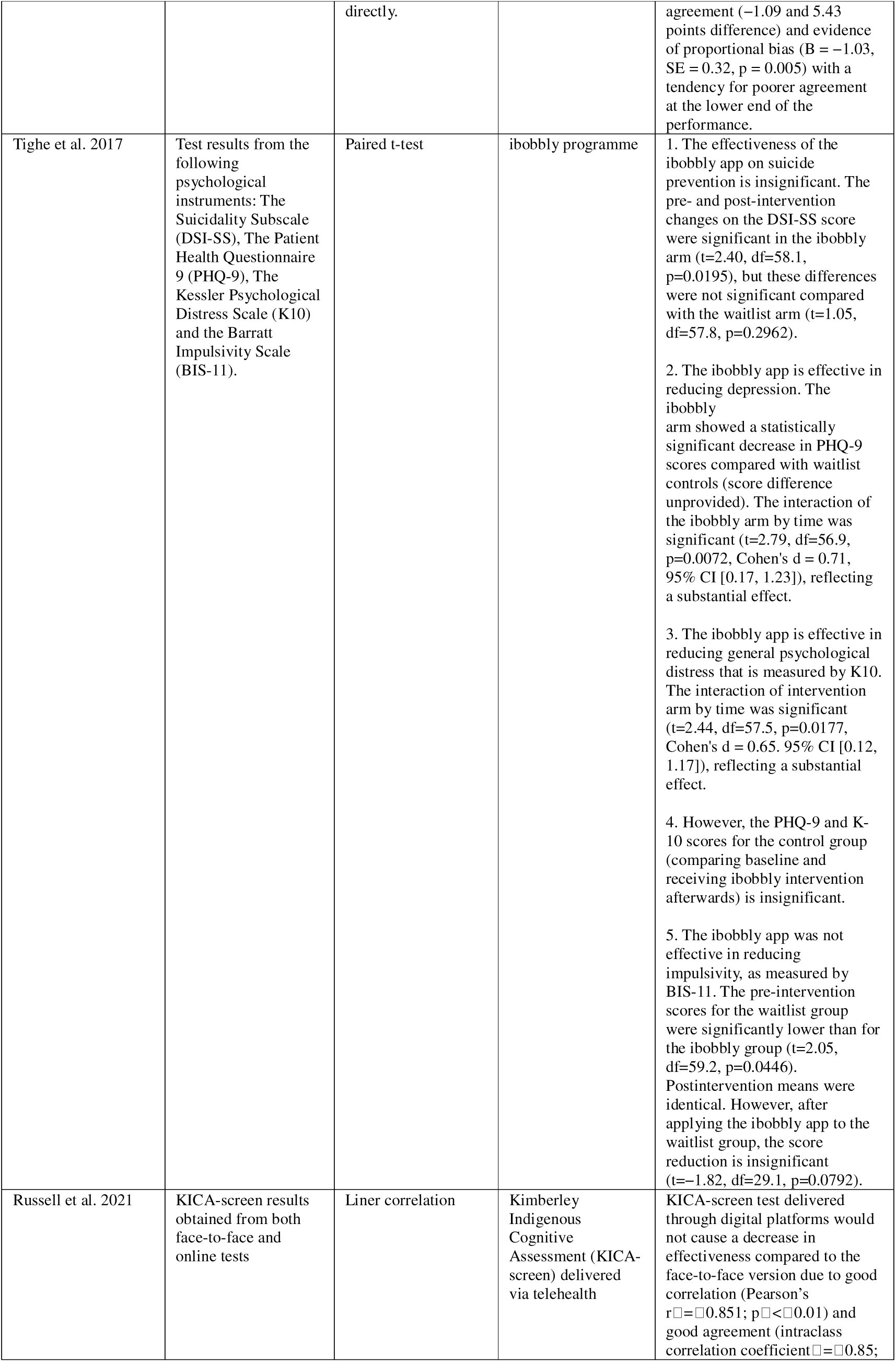
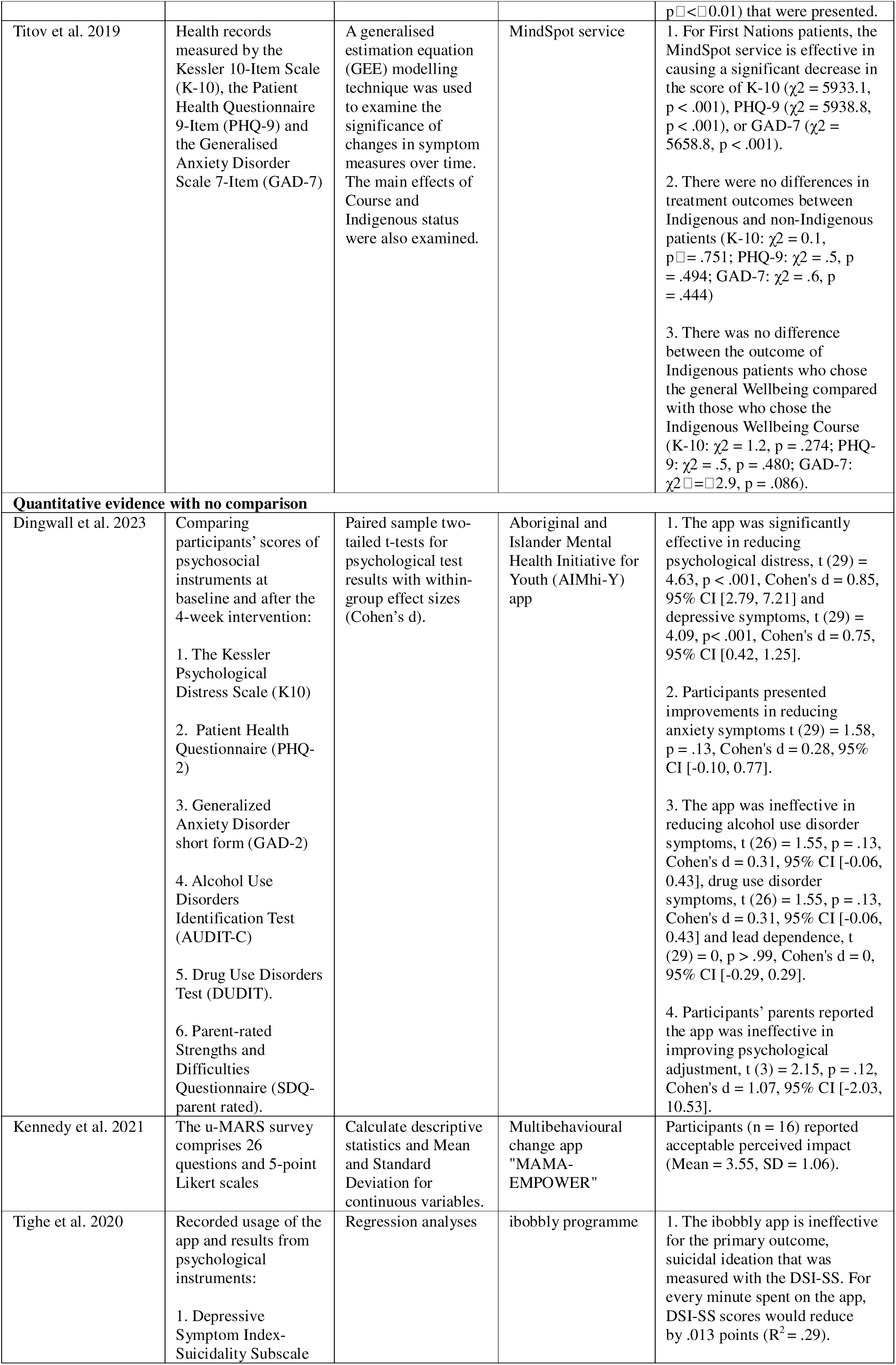
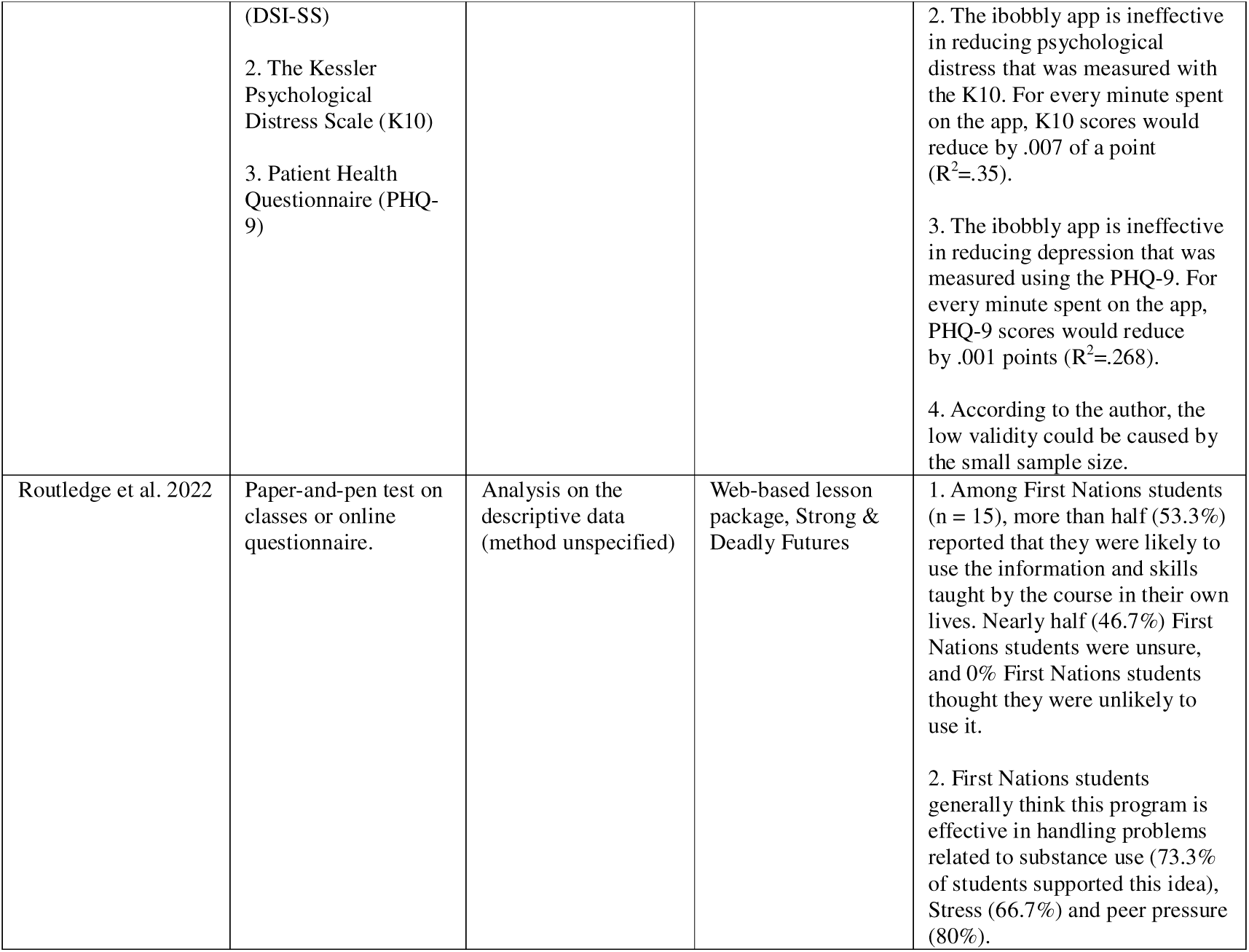
Overview of the findings in relation to the effectiveness of DMH services.

#### 2.1 Qualitative evidence

Six studies employed qualitative study designs to explore the perceived effectiveness of DMH in managing mental health conditions among First Nations Peoples (72, 77, 78, 83, 84, 86). Studies conducted by Dingwall et al. (72), Perdacher et al. (77), Povey et al. (78) applied the Youth version, Custodial version and standard version of AIMhi Stay Strong app for First Nations Peoples, respectively. The AIMhi Stay Strong app helped participants of these studies achieve mental health improvements, guided positive thinking, and supported participants in overcoming difficulties (72, 77, 78). Participants from the study of Raphiphatthana et al. (83), who mainly used the AIMhi stay strong app but expressed their perspectives on general DMH services, agreed with this effectiveness. Moreover, Effectiveness in preventing alcohol and tobacco use, and two secondary outcomes (reduced stress and peer pressure) were also reported through “Strong and Deadly” online courses aimed at preventing substance use (84). However, the participants from the study of Povey et al. (78) emphasised the effectiveness of DMH apps for people with minor mental health conditions. For another app they used, the suicide prevention ibobbly app, was reported to be insignificant due to suicide being a severe mental health condition (78). Participants from another study on the ibobbly app agreed that for “vulnerable people with strong emotions”, the effectiveness of an app could be limited (85).

#### 2.2. Quantitative evidence

There were eight studies that reported quantitatively measured effectiveness (66–68, 72, 75, 76, 85, 86). This means mental health outcomes caused by DMH services are quantified and measured by instruments and scales.

#### 2.3. Quantitative evidence without comparison

In total, four studies reported the effectiveness of DMH in managing mental health conditions among First Nations People without comparing it to typical services or other control groups (72, 75, 84, 85). Kennedy et al. (75) found that the multi-behavioural change app “MAMA-EMPOWER” was effective for the First Nations mothers due to perceived mental health impact on substance use and psychological distress, and was acceptable (3.55 average impact score in a 5-point Likert scale). Meanwhile, for a Web-based lesson package, Strong & Deadly Futures, that aimed for improving mental well-being and substance use prevention, Routledge et al. (84) found it was effective in 53.3% of First Nations students, who were likely to use the knowledge they obtained from the course, and none participants reported they would be unlikely to apply that knowledge. Moreover, 73.3%, 66.7% and 80% of First Nations students think this program is effective in managing substance use, stress and peer pressure, respectively. However, Dingwall et al. (72) found mixed effectiveness of the AIMhi-Y app that aimed to provide general mental health support for First Nations youth. More specifically, this app was effective in achieving its main objectives: reducing psychological distress and depressive symptoms (t [29] = 4.63, Cohen’s d = 0.85; t [29] = 4.09, Cohen’s d = 0.75, respectively), provided improvements in reducing anxiety symptoms (but without statistical significance), but was ineffective in managing substance use disorders, lead dependence and improving psychological adjustment. Furthermore, Tighe et al. (85) found the ibobbly app was ineffective in achieving its primary outcome: managing the suicidal ideation, and the ineffectiveness was also found in reducing psychological distress and depression. However, all indicators presented a trend of improving, according to the author, the low statistical significance could be caused by the small sample size.

#### 2.4. Quantitative evidence with comparison

Five studies examined the effectiveness of DMH services in managing (67, 86) and assessing (66, 68, 76) mental health conditions by comparing the effectiveness of DMH services with the typical mental health services (66, 68, 76), no-intervention waitlist groups (86), or comparing the effectiveness of DMH within First Nations- and Non-First Nations peoples (67).

The Grog survey app (76), which was developed for assessing substance abuse among First Nations people, was found to be effective in detecting alcohol consumption level, the overall agreement between the app and clinical interviews reached 94.7%. The app was also found to be more sensitive than clinical interview in: number of standard drinks per drinking occasion (median difference = 1.6); greatest number of drinks consumed on a single occasion (median difference = 0.8); median of reported drinking occasions (median difference = 3); average daily consumption (median difference = 0.4); classified risky drinkers (2.3% and 4.5% more detected in male and female, respectively); and the report (17.0% vs 7.2%) and prediction (p = .02 vs p = .44 in Steiger’s z-test) of tremors.

For the telehealth version of Mini Mental State Examination (TMMSE) and Kimberly Indigenous Cognitive Examination short form (TKICA-screen) (68), which tools were used for cognitive screening, compared to their typical version, the median of TMMSE scores is slightly lower than MMSE (median difference = 4.5), the agreement measured by correlation was moderate (r_s_ = 0.33, p = .2), but with an unacceptable limits of agreement, means the effectiveness may not significantly reduce in TMMSE. However, the TKICA-screen presented a significant effectiveness loss compared to the KICA-screen. For instance, there was a significant mean difference of 2.17 and evidence of proportional bias (B = −1.03, SE = 0.32, p = .005) with a tendency for poorer agreement at the lower end of the performance. Nevertheless, Russel et al. (66) also test the KICA-screen that was delivered through digital platforms, was as effective as the face-to-face version due to statistically significant correlation (Pearson’s rLJ=LJ0.851; pLJ<LJ.01) and agreement (intraclass correlation coefficientLJ=LJ0.85; p < .01) between two versions. Titov et al. (67) found that the MindSpot service that aimed to provide general mental health support was effective in managing psychological distress, depression and anxiety due to statistically significant K-10, PHQ9 and GAD-7 score decrease (χ2 = 5933.1, 5938.8, 5658.8, respectively). As a DMH service designed for all Australians, there were no statistically significant differences in either treatment outcomes between First Nations and non-First Nations patients, nor between First Nations patients who chose the general wellbeing course and the First Nations wellbeing course.

Tighe et al. (86) tested the ibobbly app that aimed for suicide prevention and set a waitlist group as a control group. The effectiveness of this app is insignificant on its primary outcome: suicide prevention, causing significant improvements on the DSI-SS score for the intervention group (t (58.1) = 2.40), but these differences were not significant compared with the waitlist arm. The ibobbly app is effective in managing depression due to the significant score decrease in the intervention group compared to the waitlist group and significant reduction of scores within the ibobbly arm by time (t [56.9] = 2.79, p =.0072, d = 0.71). Similar effectiveness was also presented in managing psychological distress (t [57.5] = 2.44, p = 0.0177, d = 0.65). However, when the control group started using the app, the effectiveness in managing depression and psychological distress was all insignificant. In terms of impulsivity management, ineffectiveness was present in all tests.

### 3. Determinants of DMH services implementation

Overall, this review found determinants that affect the DMH services implementation, and some determinants could either become facilitators or barriers. See Table 3 for extracted themes related to the determinants of DMH services implementation.

**Table 3.** Theme and subthemes of extracted studies.

| Subthemes | Supporting evidence |
| --- | --- |
| **Theme 1-Organisational factor within DMH service providers.** |  |
| The supportive structure and managers within the organisation of DMH service providers facilitated DMH service uptakes. | • Supportive structures or working culture within organisations facilitated resource allocation or staff training for DMH services uptake. (Puszka et al. 2016, Raphiphatthana et al. 2020a, Raphiphatthana et al. 2020b)<br>• Enthusiastic managers provide direct support, which increases the willingness of staff and the organisation itself for DMH service uptake. (Bennett-Levy et al. 2017, Raphiphatthana et al. 2020b) |
| Policy and structural obstacles, and a lack of resources with DMH service providers organisations create barriers for DMH service uptake. | • Procedural and administrative obstacles and obstructive IT policies limited the access of staff to technologies and DMH services and created failures in digital resource allocation. (Bennett-Levy et al. 2017, Brown et al. 2020, Raphiphatthana et al. 2020a, Raphiphatthana et al. 2020b)<br>• High workloads within the workplace and a lack of digital resources resulted in a lack of DMH training for DMH service providers. (Bennett-Levy et al. 2017, Raphiphatthana et al. 2020a, Raphiphatthana et al. 2020b) |
| **Theme 2-Cultural appropriateness of DMH services.** |  |
| The DMH services could be culturally appropriate due to cultural safety designs and culturally relevant content in DMH services, empowerment, which increased the willingness of First Nations Peoples to use DMH services. | • Overall cultural safety reported by participants (Perdacher et al. 2022).<br>• DMH services empower clients to record their own information and interact with service providers. (Puszka et al., 2016; Raphiphatthana et al., 2020b).<br>• Cultural safety designs were applied in DMH services, which created a safe environment or directly increased the acceptability and interest of clients (Dingwall et al., 2015; Fletcher et al., 2017; Povey et al., 2022; Povey et al., 2016; Snijder et al., 2021; Tighe et al., 2020). |
| A lack of cultural appropriateness created barriers for First Nations users to overcome existing negative perceptions and accept DMH services. | • The Negative perception towards the health system created barriers for First Nations Peoples to use DMH services (Brown et al. 2020).<br>• Problematic use of English within DMH services and lack of First Nations languages may cause difficulties in using DMH services (Povey et al., 2016; Snijder et al., 2021). |
| **Theme 3-First Nations People's accessibility to DMH services.** |  |
| First Nations Peoples have mixed perceptions of their accessibility to DMH services. | • First Nations clients generally appraised the high accessibility of DMH services, such as reporting the DMH services they received were accessible (Snijder et al., 2021; Titov et al., 2019).<br>• However, general accessibility-related concerns, such as the lack of services and resources and long response times, were also expressed (Brown et al., 2020). |
| The portability and flexibility of DMH services, as well as existing digital infrastructure coverage among First | • DMH services were available on multiple digital platforms (Povey et al., 2016). |
| Nations communities, could facilitate the implementation of DMH services. | • A high coverage of digital infrastructures among communities was presented, which facilitated the uptake of DMH services (Brown et al., 2020; Povey et al., 2020).<br>• DMH services can be operated on portable devices, which creates unique advantages in accessibility (Puszka et al., 2016; Tighe et al., 2020). |
| However, many First Nations peoples also reported limited access to appropriate digital infrastructures, resulting in low accessibility to DMH services. | • In some First Nations communities, the lack of digital infrastructures and devices resulted in difficulties in the implementation of DMH services (Dingwall et al., 2015; Povey et al., 2020; Puszka et al., 2016; Raphiphatthana et al., 2020a; Raphiphatthana et al., 2020b).<br>• Even in special environments such as prisons, ss to digital devices and some specific functions of DMH services, such as access to images, were limited (Perdacher et al., 2022).<br>• In remote areas, a lack of digital infrastructures created barriers for First Nations residents to implement DMH services (Povey et al., 2016; Puszka et al., 2016). |
| Free DMH apps are highly accessible, while those with download fees limit clients' access. | • Apps with high accessibility were preferred, such as free-to-download apps that can operate on different platforms (Povey et al., 2022).<br>• The cost of e-mental health apps negatively influenced their accessibility (Povey et al., 2016). |
| DMH services that do not have First Nations Language versions, creating accessibility barriers for First Nations peoples whose first language is not English. | • The lack of DMH services in First Nations languages created accessibility barriers (Dingwall et al., 2015; Povey et al. 2016; Puszka et al. 2016) |
| **Theme 4-DMH services' ability to promote engagement.** |  |
| The DMH services can promote engagement between clients and service providers by breaking the power imbalance and then creating facilitating effects. | • DMH services could help establish positive relationships between healthcare providers and clients, eventually creating positive experiences for clients or increasing First Nations clients' willingness to use those services (Dingwall et al., 2015; Puszka et al., 2016; Perdacher et al., 2022). |
| **Theme 5-The ability of DMH services to cover a wide range of settings and First Nations population groups.** |  |
| The DMH services could cover different types of clients, topics and reference periods. | • High applicability across different age groups and mental health conditions (Dingwall et al. 2015).<br>• DMH service could cover sensitive topics due to its nature of indirect engagement (Dingwall et al. 2015, Puszka et al. 2016).<br>• Compared to typical face-to-face mental health services, DMH service could cover longer reference period on substance use topic (Lee et al. 2019). |
| The DMH services failed to cover some severe mental health conditions, which was considered a barrier. | • The dMH services may not be able to cover severe mental health conditions, such as suicide (Povey et al. 2016; Puszka et al. 2016). |
| **Theme 5-The acceptability of First Nations Peoples to DMH services** |  |
| Overall, the acceptability of First Nations Peoples to DMH services was perceived as high. | • Satisfaction and optimism towards DMH services and willingness to use DMH services to seek help were presented by First Nations clients, indicating high acceptability (Dingwall et al. 2023, Dingwall et al. 2015, Perdacher et al. 2022, Kennedy et al. 2021, Povey et al. 2016, Povey et al. 2020, Routledge et al. 2022, Tighe et al. 2020, Titov et al. 2019).<br>• However, First Nations also expressed their idea of not expecting to receive help from a DMH service (Ma et al. 2022). |
| A good visual design can facilitate participants' acceptance of DMH services, while visual design problems are considered barriers. | • Clients reported the DMH service they received had an attractive or engaging visual design, which increased their acceptability of DMH services (Dingwall et al. 2015, Dingwall et al. 2023, Perdacher et al. 2022, Kennedy et al. 2021, Puszka et al. 2016). |
|  | • While some visual design problems also raised the attention of clients, such as high text density (Fletcher et al. 2017). |
| The unique nature of DMH services has high acceptability among First Nations Peoples. | • First Nations clients preferred the web-based, client-led and clinician-supported nature of DMH services (Perdacher et al., 2022, Povey et al., 2016, Snijder et al., 2021). |
| Attractive and useful content included in DMH services facilitated First Nations Peoples to accept DMH services. | • First Nations clients perceived mental health resources in DMH services are generally interactive and useful (Kennedy et al. 2021, Raphiphatthana 2020b).<br>• Embedded media forms content, such as games, videos and animation, were especially preferred by First Nations clients (Dingwall et al. 2023, Povey et al. 2016).<br>• Tailored content for First Nations students was welcomed (Snijder et al. 2021). |
| Inadequate, repetitive and less engaging content included in DMH services were considered barriers. | • Inadequate and repeating content in DMH services reduced the acceptability of DMH services (Dingwall et al. 2023, Tighe et al. 2020, Kennedy et al. 2021). |
| The unique functions of DMH services increase their acceptability among First Nations clients. | • The unique functions of DMH services, such as text messages sent as reminders or data capture functions, are acceptable for First Nations clients and prompt clients to use those services (Dingwall et al. 2023, Dingwall et al. 2015, Perdacher et al. 2022, Puszka et al. 2016). |
| Some functions are not attractive enough or hard to be used, resulting in lower acceptability of DMH services. | • However, some functions of DMH services were ignored, could hardly be used or could not fit in special contexts, such as in prison (Dingwall et al. 2023, Perdacher et al. 2022; Kennedy et al. 2021; Povey et al. 2016). |
| The DMH services with data safety have high acceptability, but the concern about data safety is perceived as a barrier. | • DMH service apps with high privacy and data safety are preferred (Povey et al. 2022).<br>• Some service providers and clients also consider data unsafety as a potential risk and barrier (Puszka et al. 2016, Brown et al. 2020). |
| **Theme 6-The ability of DMH services to integrate with existing contexts.** |  |
| Some DMH services were hard to integrate with existing contexts. | • Some service providers felt it challenging to integrate DMH service into their routine care because DMH services may find it hard to fit group clients, preexisting practices and local community status (Raphiphatthana et al. 2020a, Raphiphatthana et al. 2020b). |
| **Theme 7-The digital literacy of Clients and Service providers** |  |
| Clients and service providers who have high digital literacy facilitated the uptake of DMH services. | • Young clients presented especially high digital literacy and willingness to receive DMH services (Brown et al. 2020, Raphiphatthana et al. 2020b).<br>• Based on their own enthusiasm for using DMH services or specific DMH training, some service providers obtained high DMH literacy (Bennett-Levy et al. 2017, Brown et al. 2020). |
| Clients who do not have high digital literacy may face challenges when operating DMH services. | • Some clients report that the DMH app is difficult to operate (Fletcher et al. 2017, Povey et al. 2016, Puszka et al. 2016).<br>• Relatively lower digital literacy resulted in lower awareness to the DMH services (Dingwall et al. 2023, Povey et al. 2016; Raphiphatthana et al. 2020b). |
| Service providers who do not have high digital literacy may face challenges when operating or providing DMH services. | • Low digital literacy of service providers leads to negative perceptions, including over-estimate the complexity of technology or think DMH will distract them from their current job (Bennett-Levy et al., 2017; Brown et al., 2020; Puszka et al., 2016).<br>• Inadequate digital literacy among service providers, creating challenges in maintaining and using DMH services (Brown et al. 2020, Raphiphatthana et al. 2020a, Raphiphatthana et al. 2020b).<br>• Overall, the digital literacy level among service providers is highly heterogeneous (Puszka et al. 2016, Raphiphatthana et al. 2020a). |
| **Theme 8-The efficiency of DMH services** |  |
| DMH services have high efficiency and considered | • Service providers think DMH services can reduce the time cost of |
| timesaving. | recording client notes and waiting for data transmission, and these are advantages on data recording and management compared to typical paper format services (Puszka et al. 2016, Raphiphatthana et al. 2020b). |

#### 3.1 Organisational factors

Organisational factors, which were factors related to the administration and management of the organisation (usually service providers), could either facilitate or create barriers to implementing DMH services.

Four studies asked stakeholders and service providers to generally comment on DMH services and mentioned how organisational factors create positive effects (70, 81–83). The supportive structure within the organisations was a significant facilitator; this includes the regular supervision and review of DMH service usage (81), priority in resource, time and training allocation (82), or the work culture that welcomes DMH service implementation (83). Direct support from managers interested in DMH within the organisation was also considered to facilitate the uptake of DMH services (83) by increasing the opportunity and interest of staff in using DMH (70).

Also, four studies asked mental health service providers about their general opinion on the general DMH area (70, 82, 83) and applying DMH services in the suicide prevention area (71) reported the organisational factors that created negative effects. The lack of organisational support (83), administrative obstacles and obstructive IT policies (70, 82) within the workplace, and work restrictions (71) are the first category of negative organisational factors. The lack of training resources within organisations (82) and time allocated to cope with the extra workload brought on by the implementation of DMH (70, 82, 83) also creates organisational barriers.

#### 3.2 Cultural appropriateness

Similarly, high cultural appropriateness within DMH services was perceived as facilitators; in contrast, low cultural appropriateness would become barriers to the DMH services.

There were ten studies that reported high cultural appropriateness in DMH services (73–75, 77–79, 81, 84, 85, 87). The clients of the AIMhi Stay Strong App, which aimed to improve general mental health (73) and the custodial version of the Stay Strong App (77), expressed their overall satisfaction with the apps’ cultural safety. The clients of DMH services that aimed to improve general mental health, including the “SMS4dads” smartphone program and the “Stayin’ on Track” website (74) and AIMhi-Y app (79), and suicide prevention programme Ibobbly (85) expressed their satisfaction with the culturally appropriate visual and content design embedded in those services or simply expressed their preference (79). Notably, Both the AIMhi Stay Strong app and the Ibobbly program could create a private space to help clients avoid being judged and overcome the “shame” that First Nations clients may have (78, 85). Stakeholders and clients commented on the DMH innovations emphasised the empowerment of clients brought by some DMH services because they allowed clients to record their own information or have empowering content (75, 81).

Notably, the cultural appropriateness of the web-based substance uses prevention lesson “Strong & Deadly Futures”, which was designed for the general population, was praised. This was not only due to the culturally safe content design (First Nations characters in the story) (87), but also because it provided a fair environment for both First Nations and Non-First Nations students to be equally receptive to this program (84).

Three studies that discussed the negative aspects of cultural appropriateness within DMH services (71, 78, 87). Some First Nations health workers assumed digital suicide prevention services could be culturally inappropriate because of their perception of the current culturally unsafe mental health system (71). The clients of the AIMhi Stay Strong iPad app and the ibobbly suicide prevention app also felt uncertain about whether those apps could help them overcome the past trauma brought by the colonisation history (78). In addition, students of “Strong & Deadly Futures” online lessons expressed mixed responses towards the “Aboriginal English” within this DMH service, some of them felt uncomfortable about those words, but the other students did not (87).

#### 3.3 Accessibility

The extent to which First Nations peoples can access DMH services is another determinant of the implementation of DMH services. Two studies generally appraised the accessibility of DMH services (67, 87). For instance, some clients suggested the online clinic MindSpot could deliver service when local face-to-face mental health services are unavailable (67). The online substance use prevention course, Strong & Deadly Futures, has overall high accessibility for teachers or students with different ability levels (87). However, some First Nations peoples also expressed worry about the digital suicide prevention services due to general accessibility concerns, including poor communication systems, under-resourced nature, poor response times, and lack of services (71).

Five studies discussed the positive impact of digital infrastructures in First Nations communities (71, 78, 80, 81, 85). The AIMhi app and the iBobbly app were considered to have high accessibility because the apps can be operated on multiple devices, including tablet devices and smartphones (78). Community members and Service providers comment on applying DMH services in general mental health (80) and suicide prevention areas (71) think digital infrastructures, such as phones, are common in First Nations communities in regional or non-regional areas. Furthermore, the portability and flexibility of the ibobbly app (85) and most of the DMH services (81) facilitated the clients’ access to those services across different places, times and geographical remoteness.

In contrast, the clients or service providers who used the AIMhi app (73), the AIMhi app and the iBobbly app (78) or generally commenting on DMH services (81–83) reported they could not access appropriate digital infrastructure, including digital devices and networks, due to the low availability, which resulted in low access to DMH services, especially in special settings such as remote communities (78, 81) and prisons (77).

The cost of the app (download fee) was another ambivalent subtheme under accessibility. For example, the preference for free-to-download apps was expressed by young clients during the co-design process of the AIMhi-Y app (79), while the clients of both the AIMhi Stay Strong iPad app and the ibobbly suicide prevention app reported the download fee of apps would negatively impact their access to those services (78).

Three studies mentioned language barriers inhibiting First Nations peoples from accessing DMH services (73, 78, 81). The clients of the AIMhi Stay Strong App (73) and both the AIMhi App and the iBobbly app (78) reported that those apps do not have First Nations Language versions. This resulted in difficulties in using apps when clients could not understand some English words (such as “resilience”) (78), and as a group of stakeholders commented, language barriers limited First Nations peoples whose first language is not English to access many DMH services (81).

#### 3.4 Integration

The integration of DMH services with existing practices was generally perceived as a barrier in the included studies. More specifically, some service providers in two studies commented on applying DMH in general mental health areas (82, 83) reported difficulties in integrating DMH into the usual practices when facing non-one-to-one settings, such as facing a group of clients (82) or need to fit the existing practices and local community status (83).

#### 3.5 Engagement (between service providers and clients)

According to the three included articles (73, 77, 81), DMH services’ feature of promoting engagement between clients and service providers was welcomed in general mental health areas. As some service providers indicated, the AIMhi app was able to overcome barriers (such as power imbalance) between healthcare providers and clients, therefore establishing positive relationships between them (73). Also, stakeholders expanded this result to other DMH tools that could provide indirect engagements when solving sensitive issues and supported the building of positive relationships (81). Notably, the improved rapport was observed in a special environment: the custodial context by applying the custodial version of the Stay Strong App (77).

#### 3.6 Coverage

The coverage refers to the ability of DMH services to cover different types of clients and various mental health conditions. Three studies indicated that the high coverage of DMH services facilitated the implementation (73, 76, 81). Some service providers introduced that the AIMhi App can cover different types of clients in different age groups, with different mental health issues, or who were hard to engage with when providing general mental health supports (73). Also, clients of the AIMhi App (73) and service providers providing general opinions on DMH (81) indicated the ability of DMH services to cover sensitive mental health topics. Furthermore, compared with typical clinical interviews, the Grog Survey App aimed to investigate alcohol dependence status can cover a longer reference period (76).

However, two studies discussed client feedback on both the AIMhi app and the iBobbly app (78), and service providers’ opinions on DMH (81), suggested that DMH services may not be able to cover severe mental health conditions, such as suicide.

#### 3.7 Acceptability

Acceptability refers to the extent to which participants can accept the DMH services. Ten studies mentioned the overall acceptability of DMH services (65, 67, 72, 73, 75, 77, 78, 80, 84, 85). Except for the DMH service in one study, lifeline Australia was low in acceptance because participants reported low expectations to receive suicide prevention support from this app (without specifying reasons) (65), the other nine studies were all reported as generally acceptable by participants’ acceptable usage, marks on the overall quality and satisfaction, and comments relevant to this theme. These DMH services include the AIMhi Stay Strong App (73), its custodial version (77) and its youth version (72), the “MAMA-EMPOWER” app (75), both the AIMhi app and the iBobbly app (78), web-based lessons “Strong & Deadly Futures” (84), iBobbly app (85), online clinic MindSpot (67), and some participants’ general comments on DMH services (80).

The nature of DMH services was reported as acceptable by three studies (77, 78, 87). The custodial version of the AIMhi App was appraised for its client-led nature (77), the AIMhi app and the ibobbly suicide prevention app were praised for their clinician-supported nature that could overcome mental health literacy barrier (78), and the web-based nature preferred by students of web lessons “Strong & Deadly Futures” (87).

Two studies mentioned the relevance of DMH services (75, 84). Some clients of the Multibehavioural change app “MAMA-EMPOWER” thought this app was relevant to their issue or situation (75). However, the substance use content of the “MAMA-EMPOWER” app (75) and online lessons, “Strong & Deadly Futures” (84) were considered irrelevant by mothers who did not consume substances (75) or students who thought they were too young to discuss this topic (84).

Four studies indicated some unique digital functions of DMH services were acceptable (72, 73, 77, 81). For example, some functions that were highlighted as useful, including the text messages function of the AIMhi-Y app (72), the function of ensuring service consistency across the practice setting of the AIMhi app (73), the goal-setting functions of the custodian version of the Stay Strong App (77), or the data capture functions reported by service providers generally discussed DMH services (81).

However, some functions did not create enough facilitating effects, including the “stories” or the “get help page” in the AIMhi-Y app (72) that did not raise the attention of the clients or the lack of personalisation of avatar functions that were tailored to the special custodial context of the AIMhi app custodial version (77). Also, some of the clients of the Multibehavioural change app “MAMA-EMPOWER” (75) and both the AIMhi app and the ibobbly app reported some functions or the whole app was hard to use (78).

Five studies mentioned the subtheme that acceptable content of DMH services were facilitators (72, 75, 78, 83, 87). The overall content in DMH services (83) and the Multibehavioural change app “MAMA-EMPOWER” (75) were positively commented on because the participants thought the content was useful and engaging. The embedded media content, including games, videos, animation and graphics in the AIMhi-Y app (72) and both the AIMhi app and the ibobbly app (78) were generally welcomed by the participants. Notably, the content specifically designed for First Nations students in web-based substance dependence lessons, “Strong & Deadly Futures”, was considered informative (87).

However, three studies mentioned the barrier caused by insufficient content in DMH services (72, 75, 85). As the clients of the AIMhi-Y app (72), ibobbly programme (85) and the Multibehavioural change app “MAMA-EMPOWER” (75) mentioned, the content of those DMH services was inadequate to provide enough information, and the contents were highly repetitive after the first use (72, 85).

There were six studies that mentioned the subtheme of acceptability towards the visual design of DMH services (72–75, 77, 81). Except for the clients of the “SMS4dads” smartphone mental health support program and the “Stayin’ on Track” website (74) reported there was a lack of attractive visual design and the current design was too text-dense; other First Nations peoples positively commented on the visual design of DMH services. This includes the AIMhi APP (73), its youth version (72), and the custodial version (77), which were reported to have appropriate and attractive visual designs. Similarly, the colour and overall visual design of the Multibehavioural change app “MAMA-EMPOWER” was acceptable for First Nations mothers (75). Besides, stakeholders who were commenting on DMH services emphasised the facilitating effects of easy-to-navigate or self-explanatory visual designs (81).

Three studies mentioned data safety and privacy as a subtheme (71, 79, 81). Some youth clients mentioned their preference for DMH services with data safety during the co-design process of the AIMhi-Y app (79). However, the concerns about data safety in applying DMH services in general mental health (81) and suicide prevention (71) areas were perceived as barriers to DMH services implementation.

#### 3.8 Digital literacy

Three studies discovered how high digital literacy has facilitated the implementation of DMH services (70, 71, 83). Higher digital literacy levels among young First Nations clients, which resulted in their higher acceptability towards DMH, were specifically emphasised in two studies asking the general opinion of participants on the DMH services (71, 83). Some suicide prevention gatekeepers expressed their preference for using DMH platforms (such as Facebook) to provide services (71). For some service providers in general DMH areas, digital literacy improved after they received DMH training, resulting in higher acceptability of DMH services (70).

However, low digital literacy could become a barrier for clients (72, 74, 78, 81, 83) or service providers (70, 71, 81–83) in utilising DMH services. For clients, difficulties in operating DMH services apps due to not having enough digital literacy were observed from First Nations fathers who used the “SMS4dads” smartphone program and the “Stayin’ on Track” website for mental health help (74), and clients of AIMhi app and ibobbly app (78), and the general opinion of participants on DMH services (81). Moreover, as the clients of the AIMhi App and ibobbly app reported, First Nations community members may have low awareness of DMH tools (78). For example, both of a part of First Nations Youth clients of the AIMhi-Y app (72) and First Nations clients expressed their general opinion on DMH services (83) frequently ignore or forget to use the DMH services.

For service providers, heterogeneous overall digital or mental health literacy levels were observed among service providers working in the general mental health areas (81, 82). This could be presented as some “less technologically competent” suicide gatekeepers (71) or service providers commenting on the general DMH area (82, 83) perceived difficulties in using technologies (71) and operating basic functions of the DMH services (i.e. downloading the DMH apps) (82, 83). Also, existing negative perceptions of DMH services were considered another type of barrier. This includes participants who expressed their opinion on applying DMH in general mental health (70, 81) and suicide prevention area (71) and reported their existing negative perception of DMH services, including worrying about the complexity and novelty of DMH (70), consider DMH was potentially less efficient and would bring extra workload (81) and think using DMH would distract them from working (71).

#### 3.9 Efficiency

Two groups of service providers provided opinions on general DMH services (81, 83) and indicated the improved efficiency brought by DMH services. More specifically, DMH services could reduce the time cost of recording client notes and waiting for data transmission (81), then became advantages in data recording and management compared to typical paper format services (83).

## Discussion

### 1. Principal findings

This is the first review to synthesise evidence on the effectiveness, facilitators and barriers of DMHs for First Nations people in Australia. Our study found that DMH was effective in assessing mental health conditions, monitoring mental health status, and delivering mental health education and had the potential to improve mental health conditions among First Nations Australians. However, its effectiveness on severe mental health conditions remained limited. Additionally, this study also identified determinants that could facilitate or inhibit the uptake of DMHs among First Nations people in Australia.

### 2. Effectiveness of DMHs

The effectiveness of DMHs for First Nations Australians varied across different mental health outcomes and intervention types. Consistent with a previous government report (89) and similar studies (90–92), qualitative studies included in the current review suggest that DMH interventions can support mental health improvement, increase self-reflection, and enhance self-confidence and empowerment (72, 77, 78, 83–85). Considering most included DMH services applied culturally adapted designs, these findings align with broader evidence that culturally adapted DMH tools can provide valuable support for First Nations Peoples’ mental health (4, 11, 93). However, concerns remain about the adequacy of digital interventions for severe mental health conditions, particularly suicide prevention, where some participants expressed doubts about their effectiveness beyond mild to moderate distress (78, 85, 86). This highlights the ongoing challenge of delivering digital suicide prevention interventions effectively in First Nations communities, as also noted in previous research (57).

Moreover, evidence supports the effectiveness of DMH interventions for substance use. The Grog Survey App accurately identified at-risk drinkers with high sensitivity and reliability (76) and was reported to be effective by suggesting its potential application in screening and risk assessment. The web-based lesson package, Strong & Deadly Futures, was perceived to be effective in preventing substance use (84). However, the AIMhi-Y app was ineffective in reducing alcohol and drug use symptoms (72), indicating that while DMH tools could be valuable for monitoring and early intervention, they might not provide sufficient therapeutic impact for substance use disorders without additional face-to-face support. This aligns with previous research on First Nations peoples in the U.S. that received insignificant therapeutic outcomes (94). The use of DMH interventions for cognitive assessment also yielded mixed results (66, 68). These findings highlight the need for careful adaptation of cognitive assessments for telehealth delivery, as the telehealth version of the test reduced the visual cues of tests (68) and resulted in insignificant results, but the same test that was delivered through video conferencing obtained acceptable effectiveness compared with typical in-person services (66). Finally, our engagement and user perception findings indicate that digital mental health education and behavioural change interventions can be beneficial but require further refinement. For instance, the Strong & Deadly Futures program was generally well-received, with 73.3% of First Nations students reporting that it was effective in addressing substance use (84). However, a substantial proportion of students remained unsure about applying the information in their own lives. This suggests that while digital education programs can improve mental health literacy, their impact on long-term behavioural change may be limited without additional reinforcement, as noted in previous research (95).

### 3. Determinants of DMH uptake among First Nations people in Australia

The current systematic review also identified determinants of (i.e., the facilitators and barriers to) the uptake of DMHs among First Nations people in Australia. Our study found that strong leadership is critical in DMH adoption, highlighting the crucial role of organisation support in digital health uptake among First Nations Australians (70, 71, 81–83). Leadership drives the strategic vision, fosters innovation, and ensures resource allocation, which is essential for successful implementation (96, 97). Strong leadership also facilitates stakeholder engagement and addresses resistance to change, both key factors in digital health uptake (98). Organisational support further enhances workforce readiness and policy development, sustaining long-term adoption (99). Without these elements, DMH initiatives risk poor adoption and limited impact (70, 71, 82, 83). Additionally, culturally relevant app designs incorporating First Nations art and metaphors were found to enhance engagement (73, 74, 78, 79, 85, 87). This echoes prior research emphasising the importance of embedding cultural elements to ensure relevance and acceptance within First Nations communities (100).

First Nations youth were also found to prefer mobile apps, as they tend to be more tech-savvy and exhibit higher engagement with digital mental health interventions (72, 79, 87). This is consistent with previous research highlighting that mobile-based interventions can be particularly effective for younger populations due to their familiarity with digital platforms and preference for flexible, on-demand mental health support (101). Additionally, community-driven initiatives, such as private Facebook groups, were reported as valuable in supporting peer-to-peer learning and engagement. Social media platforms have been recognised as important tools in mental health interventions for First Nations communities, as they provide a culturally safe space for connection, knowledge-sharing, and mental health promotion (91).

Meanwhile, our study also identified several barriers to DMH adoption among First Nations people in Australia. Despite some evidence shows common ownership of digital infrastructures in some First Nations communities (71, 80), limited infrastructure and technology access remain significant challenges, particularly in remote communities where access to tablets, Wi-Fi, and reliable technology is often restricted (71, 73, 78, 80–83). It has been reported that First Nations Australians, especially those living in remote areas, are one of the most digitally excluded populations (102). This finding highlights the urgent need for targeted investments in digital infrastructure to bridge the technology gap and ensure equitable access to DMH services. To address these challenges, it could be helpful to expand community Wi-Fi programs, provide subsidised digital devices, and implement mobile-based DMH solutions that require minimal bandwidth (89). The Mob Link initiative by the Institute for Urban Indigenous Health (IUIH) is a strong example (103), as it provides culturally responsive digital health support, including assistance with telehealth and digital access, helping to reduce barriers and improve engagement with healthcare services.

Secondly, low digital literacy and privacy concerns were found to reduce trust and engagement with DMHs. Inconsistent digital literacy among both clients and practitioners has been previously identified as a key obstacle to effective DMH utilisation (70–72, 74, 78, 81–83). Delivering technology training and ongoing support could improve digital literacy and confidence in using DMH services (104, 105), then ultimately enhancing their uptake and efficacy among First Nations communities. Additionally, privacy concerns and fears of data misuse were significant deterrents to DMH uptake (71, 81). This aligns with previous research that highlights First Nations people’s concerns over data sovereignty and the potential misuse of personal health information (106). Ensuring that DMH services are designed with robust privacy protections and transparent data governance frameworks is crucial to fostering trust and engagement.

### 4. Limitation

We acknowledge that this review is subject to several limitations. First, the included studies were highly heterogeneous regarding intervention types, study designs, and outcome measures. This limits our ability to quantify overall effectiveness. Second, this review could have missed relevant grey literature.

### 5. Conclusion

This study highlights the effectiveness of DMHs in assessing, monitoring, and managing mental health conditions for First Nations Australians, though their impact on severe conditions remains limited. Key facilitators include strong leadership, culturally relevant designs, clinician-supported tools, and community-driven initiatives, whilst major barriers—such as digital exclusion, low digital literacy, and privacy concerns— highlight the need for targeted infrastructure investment, digital training, and transparent data governance. Initiatives like Mob Link by IUIH demonstrate how culturally responsive support can enhance engagement. Future research should focus on co-designing DMHs with First Nations communities, rigorously evaluating their effectiveness, and integrating them into existing healthcare models to ensure accessibility, cultural relevance, and long-term impact.

## Supporting information

Supplementary File S1 Searching strategy

Supplementary File S2 Extracted results

Supplementary File S3 Cultural safety assessment

Supplementary File S4 MMAT assessment results

## Data Availability

All data produced in the present work are contained in the manuscript.

## Appendix A

| Patient/Population | Intervention | Comparison | Outcomes |
| --- | --- | --- | --- |
| - First Nations Peoples in Australia who have mental health conditions or are at risk of having | - Digital Mental Health Services, including:<br>- Screening, diagnosis and | - Typical non-digital mental health services | - Accurate diagnosis and screening of mental health conditions<br>- Relieve or improve symptoms of |
| mental health conditions. | treatment towards mental illness.<br>- Prevention and support for mental health issues, including suicide or self-harm behaviours and substance use. | - No comparison | mental health conditions<br>- Prevention of mental health conditions. |

| Inclusion Criteria | Exclusion Criteria | Extra Criteria |
| --- | --- | --- |
| - Be written in English<br>- Studies on First Nations Peoples in Australia<br>- Studies on DMH services<br>- Studies on digital health services have a purpose other than mental health but contain DMH services as their components.<br>- Studies on DMH services provided for the general population but specified the data of First Nations Peoples.<br>- Studies on users' or service providers' experience of using DMH services.<br>- Studies on determinants affecting the use of DMH services | - Studies on First Nations Peoples outside of Australia.<br>- Studies on Non-First Nations People.<br>- Studies that have not researched digital mental health services.<br>- Studies irrelevant to the determinants affecting the use of DMH services<br>- Review articles, conference papers, research protocols, and other non-primary research studies.<br>- Full text unavailable online. | - No specific limit to the publication dates of studies (DMH is a relatively new area)<br>- No specific limit on the age of participants<br>- No limit on the nation groups of First Nations Peoples in Australia |

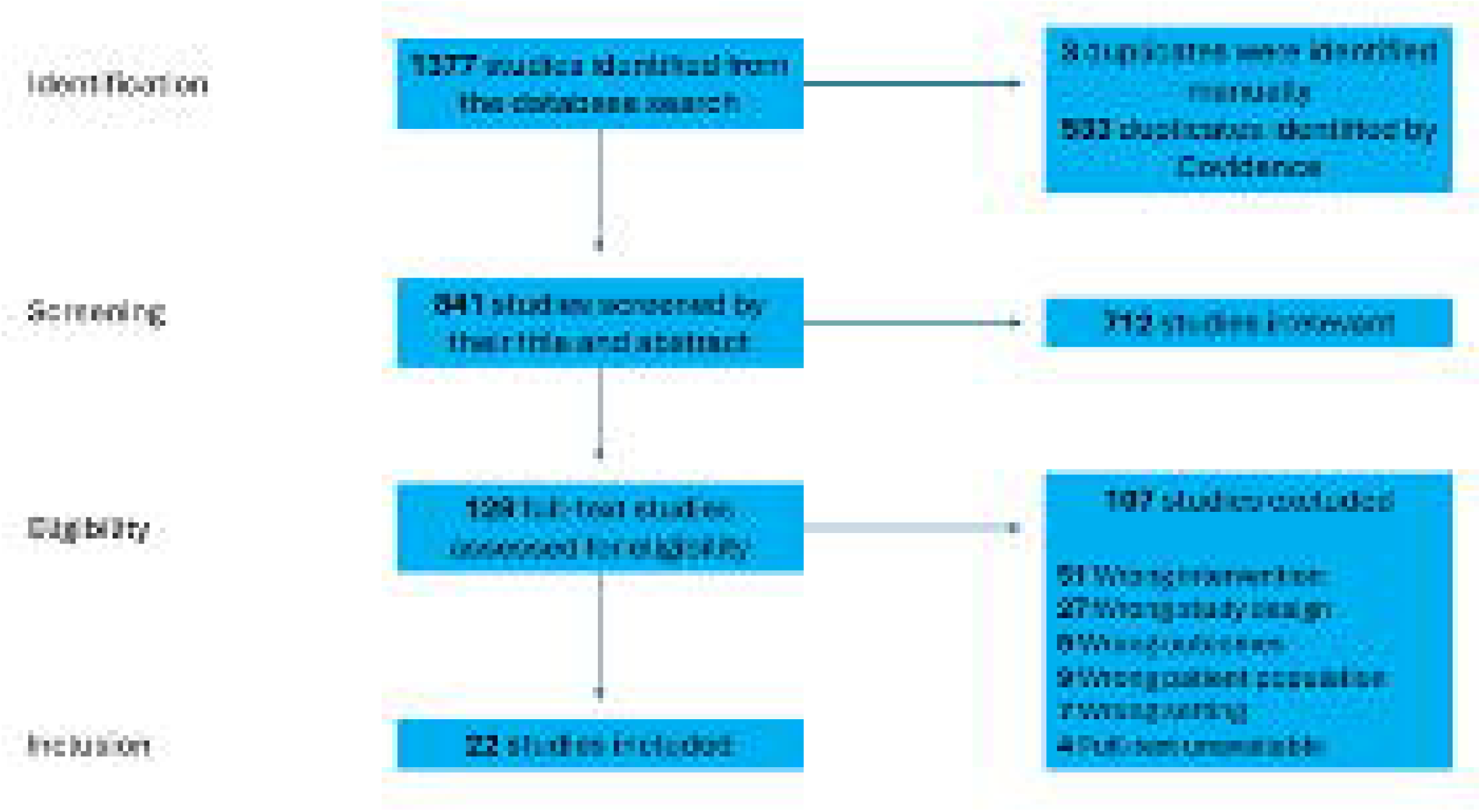

