## Supplementary File S1 Searching strategy for "Effectiveness, facilitators and barriers of digital mental health services for First Nations Peoples in Australia: A systematic review"

Search terms:

First Nations Peoples:

Indigenous OR Aboriginal OR "First Nations" OR " Torres Strait Islander" OR ATSI

Mental health:

"Mental illness" OR "Psychological disorder" OR "Mental disorder" OR "Mental health" OR "Psychotic Illness" OR “Psychiatric disorder” OR “Mental Condition

Digital Mental Health:

"Digital health" OR "Digital mental health" OR telemedicine OR telehealth OR ehealth OR e-health OR "electronic health" OR "health apps" OR mhealth OR m-health OR "mobile health" OR "online health" OR "eMental Health" OR “Mental Health Apps” OR Web-based OR mtherapy OR “Online therapy” OR “Online intervention” OR “Online Self-help”

Mental Health Conditions:

Depression OR “Major depressive disorder” OR Dysthymia OR Anxiety OR Schizophrenia OR “Bipolar disorder” OR “Eating Disorder” OR “Anorexia nervosa” OR “Bulimia nervosa” OR “Attention-deficit/hyperactivity disorder” OR ADHD OR “Autism Spectrum Disorder” OR “Conduct disorder” OR “Idiopathic developmental intellectual disability” OR Suicide OR Self-harm OR Suicidal

Australia:

Australia* OR Victoria OR Queensland OR Tasmania OR "New South Wales" OR "Western Australia" OR "Northern Territory" OR "Australian Capital Territory" OR "South Australia"
